# Insufficient fibrinolysis in COVID-19: a systematic review of thrombolysis based on meta-analysis and meta-regression

**DOI:** 10.1101/2020.09.07.20190165

**Authors:** Hong-Long Ji, Zhenlei Su, Runzhen Zhao, Andrey A. Komissarov, Guohua Yi, Shan-Lu Liu, Steven Idell, Michael A. Matthay

## Abstract

**Background:** How aberrant fibrinolysis influences the clinical progression of COVID-19 presents a clinicopathological dilemma challenging intensivists. To investigate whether abnormal fibrinolysis is a culprit or protector or both, we associated elevated plasma D-dimer with clinical variables to identify a panoramic view of the derangements of fibrinolysis that contribute to the pathogenesis of COVID-19 based on studies available in the literature.

**Methods:** We performed this systematic review based on both meta-analysis and meta-regression to compute the correlation of D-dimer at admission with clinical features of COVID-19 patients in retrospective studies or case series. We searched the databases until Aug 18, 2020, with no limitations by language. The first hits were screened, data extracted, and analyzed in duplicate. We did the random-effects meta-analyses and meta-regressions (both univariate and multivariate). D-dimer associated clinical variables and potential mechanisms were schematically reasoned and graphed.

**Findings:** Our search identified 42 observational, or retrospective, or case series from six countries (n=14,862 patients) with all races and ages from 1 to 98-year-old. The weighted mean difference of D-dimer was 0.97 μg/mL (95% CI 0.65, 1.29) between relatively mild (or healthy control) and severely affected groups with significant publication bias. Univariate meta-regression identified 58 of 106 clinical variables were associated with plasma D-dimer levels, including 3 demographics, 5 comorbidities, 22 laboratory tests, 18 organ injury biomarkers, 8 severe complications, and 2 outcomes (discharge and death). Of these, 11 readouts were negatively associated with the level of plasma D-dimer. Further, age and gender were confounding factors for the identified D-dimer associated variables. There were 22 variables independently correlated with the D-dimer level, including respiratory rate, dyspnea plasma K^+^, glucose, SpO_2_, BUN, bilirubin, ALT, AST, systolic blood pressure, and CK. We thus propose that insufficient hyperfibrinolysis (fibrinolysis is accelerated but unable to prevent adverse clinical impact for clinical deterioration COVID-19) as a peculiar mechanism.

**Interpretation:** The findings of this meta-analysis- and meta-regression-based systematic review supports elevated D-dimer as an independent predictor for mortality and severe complications. D-dimer-associated clinical variables draw a landscape integrating the aggregate effects of systemically suppressive and locally (i.e., in the lung) hyperactive derangements of fibrinolysis. D-dimer and associated clinical biomarkers and conceptually parameters could be combined for risk stratification, potentially for tracking thrombolytic therapy or alternative interventions.

**Funding:** National Institute of Health.

## Introduction

The sustained COVID-19 pandemic has oversaturated the emergency and intensive critical care resources globally. Hypercoagulability has been evidenced in most critically ill patients by elevated D-dimer and fibrin degradation products (FDP), a decrease in platelet count, an incremental increase in the prothrombin time, and a rise in fibrinogen^1-9^. Of these, patients with increased D-dimer are more vulnerable to worsened clinical consequences of COVID-19, with more severe complications, including require ICU support^1-9^.

Thromboembolism of COVID-19 patients is the fatal sequelae of hypercoagulation and fibrinolytic abnormalities. Pulmonary embolism (PE) and deep vein thrombosis (DVT) can cause respiratory failure in severely ill patients with COVID-19^10-14^. Postmortem pathology shows that small fibrinous thrombi in small pulmonary arterioles are very common. Activation of the coagulation cascade is further supported by endothelial tumefaction, pulmonary megakaryocytes in the capillaries, and endotheliitis^15-19^. Elevated D-dimer is an indicator of the activation of the fibrinolysis system and removal of clots or extravascular collections of fibrin by plasmin. Compared with the consistent coagulopathy, however, the clinical ramifications of deranged fibrinolysis are not well studied and reviewed systematically.

Increased D-dimer level has not consistently been observed by all COVID-19 clinical studies, although it is a broadly applied biomarker for prognosis and outcomes of anti-thrombosis^20^. The current explanations for the elevated D-dimer in critically ill patients are multiple, including “suppression of fibrinolysis”, “secondarily hyperactive fibrinolysis”, “consumption of fibrinolysis”, “fibrinolysis resistance”, and “fibrinolysis shutdown”^21,22^. To address the coagulopathic changes complicating COVID-19, two diametrically different therapeutic regimes are in practice: fibrinolytic (alteplase-tPA)^10,11,23-28^ and antifibrinolytic therapies (nafamostat and tranexamic acid (TXA))^29-31^. It is therefore imperative to clarify the role of pathophysiologic derangements of fibrinolysis in clinical outcomes that occur in COVID-19 patients. We therefore systematically reviewed key cohort studies and performed both meta-analyses and meta-regressions to explore the relationships between the plasma D-dimer level on admission with demographics, laboratory tests, fatal cardiopulmonary function, radiology, interventions, complications, and outcomes. As a result, we herein propose a model, namely, the insufficient hyperfibrinolysis within the inflamed organs that explains the aberrant metabolism, distribution, and excretion of fibrin(ogen) in COVID-19 patients.

## Methods

We conducted a systematic review of literature in accordance with the methods recommended in the PRISMA guidelines (**Figure S1**).

### Literature search

Two independent investigators searched the potential studies in the NCBI PubMed, EMBASE, Scopus, Web of Science, Google Scholar, and some preprint platforms, including the medRxiv, Preprint, and bioRxiv. The search strategy was (D-dimer OR fibrin OR proteolytic OR fibrinolysis OR coagulation OR thrombin OR platelet OR plasmin OR tPA OR fibrinolytic OR thrombolytic) AND (COVID-19 OR 2019-nCoV OR SARS-nCoV OR Wuhan OR SARS-CoV-2). Sorted COVID-19 and SARS-CoV-2 preprints were screened if available. The hits were limited to publication in year of 2019-2020. Studies published in some high impact journals focusing on the fibrinolysis and coagulation systems were summarized (**Table 1**). Related articles published during the preparation of this manuscript were discussed.

**Table 1.** Demographic features of analyzed 42 studies. *IQR. Mean ± S.D. # normal and D-dimer elevated group. (-) range from minimum to maximum. N/A, not available. μ AS, Asian. AF, African. CC, Caucasian. MR, mixed race, or others. Normal D-dimer range, < 0.5 μg/ml.

| Study | Journal | Region | Cases | Age (year) | Race | Male /% | D-dimer ($\mu\text{g/mL}$) | Mortality /% |
| --- | --- | --- | --- | --- | --- | --- | --- | --- |
| Akalin E, et al<sup>9</sup> | NEJM | USA | 36 | 60 (32 - 77) | AF/CC/MR | 26/72 | 1.02 (0.32 - 5.19) | 10/28 |
| Borba MG, et al<sup>102</sup> | JAMA Network | Brazil | 81 | 51.1 ± 13.9 | CC/AA/MR | 61/75.3 | N/A | 22/27.2 |
| Chen G, et al<sup>33</sup> | JCI | China | 21 | 56 (50, 65)* | Asian | 17/81 | 0.5 (0.4, 1.8)* | 4/19 |
| Chen N, et al<sup>103</sup> | Lancet | China | 99 | 55.5 ± 13.1 | Asian | 67/68 | 0.9 (0.5, 2.8)* | 11/11 |
| Chen T, et al<sup>34</sup> | BMJ | China | 274 | 62 (44, 70)* | Asian | 171/62 | 1.1 (0.5, 3.2)* | 113/41.2 |
| Cui S, et al<sup>104</sup> | J Thromb Haemost | China | 81 | 59.9 ± 14.1 | Asian | 37/46 | 3 (0 - 8.2) | 8/10 |
| Du R, et al<sup>105</sup> | ERJ | China | 179 | 56.7 ± 13.7 | Asian | 97/54.2 | 0.5 (0.3, 1.7)* | 21/11.73 |
| Du Y, et al<sup>106</sup> | AJRCCM | China | 85 | 65.8 ± 14.2 | Asian | 62/72.9 | 5.16 ± 4.68 | 81/95.29 |
| Feng Y, et al<sup>35</sup> | AJRCCM | China | 476 | 53 (40, 64)* | Asian | 271/56.9 | 0.58 (0.35, 1.48)* | 38/8 |
| Fogarty H, et al<sup>107</sup> | BJH | Ireland | 83 | 62 ± 16.3 | CC/AS/AF | 55/66.27 | 0.88 (0.74, 3.46)* | 13/15.7 |
| Han H, et al<sup>108</sup> | CCLM | China | 94 | N/A | Asian | 48/51 | 10.36 ± 25.31 | N/A |
| Helm J, et al<sup>13</sup> | ICM | France | 150 | 63 (53,71)* | CC/AA/MR | 122/ 81.3 | 2.27 (1.16, 20.0)* | 13/8.7 |
| Huang C, et al<sup>36</sup> | Lancet | China | 41 | 49 (41, 58)* | Asian | 30/73 | 0.5 (0.3, 1.3)* | 6/15 |
| Lu JT, et al<sup>8</sup> | Lancet Infect Dis | China | 577 | 55 (39,66)* | Asian | 254/44 | 0.3 (0.1, 0.7)* | 39/6.8 |
| Mo P, et al<sup>37</sup> | CID | China | 155 | 54 (42, 66)* | Asian | 86/55.5 | 0.19 (0.12, 0.36)* | 22/14.2 |
| Oxley TJ, et al<sup>32</sup> | NEJM | USA | 5 | 40.4 (33 - 49) | CC | 4/80 | 3.66 (0.05 - 13.8) | 0 |
| Panigada M, et al<sup>49</sup> | J Thromb Haemost | Italy | 24 | 56 (23 - 71) | CC | N/A | 4.88 (1.2 - 16.95) | N/A |
| Paranjpe I, et al<sup>38</sup> | medRxiv | USA | 2,199 | 65 (54 - 76)* | CC/AA/AS/MR | 1293/58.8 | 1.31 (0.74, 2.44)* | 310/14.10 |
| Qiu H, et al<sup>109</sup> | Lancet Infect Dis | China | 36 | 8.3 ± 3.5 | Asian | 23/64 | 0.29 ± 0.2 | 0 |
| Ranucci M, et al<sup>39</sup> | J Thromb Haemost | Italy | 16 | 61 (55, 65)* | CC | 15/93.75 | 2.5 (1.6, 2.8)* | 7/43.7 |
| Rentsch CH, et al<sup>40</sup> | medRxiv | USA | 585 | 66.1 (60.4,71.0)* | CC/AF/MR | 558/95.4 | N/A | 17/2.9 |
| Richard S, et al<sup>7</sup> | JAMA | USA | 5,700 | 63 (52, 75)* | AF/AS/CC/MR | 3437/60.3 | 0.44 (0.26, 0.87)* | 553/21 |
| Spiezia L, et al<sup>110</sup> | Thromb Haemost | Italy | 22 | 67 ± 8 | CC | 20/90.91 | 5.34 ± 2.10 | 1/4.55 |
| Tang N, et al<sup>111</sup> | J Thromb Haemost | China | 449 | 65.1 ± 12.0 | Asian | 268/59.69 | 1.94 (0.9, 9.44)* | 134/29.8 |
| Wang D, et al<sup>6</sup> | JAMA | China | 138 | 56 (22 - 92) | Asian | 75/54.3 | 0.20 (0.12 - 0.40) | 6/4.3 |
| Wang L, et al<sup>41</sup> | J Infect | China | 339 | 69 (65 - 76)* | Asian | 166/49 | 1.20 (0.62, 3.25)* | 65/19.2 |
| Wang Y, et al<sup>42</sup> | AJRCCM | China | 344 | 64 (52, 72)* | Asian | 179/52.0 | 1.3 (0.5, 5.0)* | 133/38.7 |
| Wang YM, et al<sup>5</sup> | Lancet | China | 158 | 66.0 (57, 73)* | Asian | 89/56 | N/A | 22/15 |
| Wang Z, et al<sup>43</sup> | CID | China | 69 | 42 (35, 62)* | Asian | 32/46 | N/A | 5/7.5 |
| Wu C, et al<sup>112</sup> | JAMA Inter Med | China | 201 | 51 (43, 60)* | Asian | 128/63.7 | 0.61 (0.35, 1.28)* | 44/21.9 |
| Wu J, et al<sup>44</sup> | CID | China | 80 | 46.1 ± 15.42 | Asian | 39/48.75 | 0.9 (0.4, 2.4)* | 0 |
| Xie J, et al<sup>3</sup> | Lancet | China | 299 | 62 (50.8, 71)* | Asian | 239/53.8 | N/A | 224/50.5 |
| Xu X, et al<sup>4</sup> | BMJ | China | 62 | 41 (32, 52)* | Asian | 36/58 | 0.2 (0.2, 0.5)* | 0 |
| Yang F, et al<sup>45</sup> | I Med Virol | China | 52 | 63 (34 - 98)* | Asian | 28 (53.8) | 1.7 (0.7, 3.3)* | 11/21.2 |
| Yang WJ, et al<sup>113</sup> | J Infect | China | 149 | 45.11 ± 13.35 | Asian | 81/54.4 | 0.22 ± 0.28 | 0 |
| Yang X, et al<sup>114</sup> | Lancet Respir Dis | China | 52 | 59.7 ± 13.3 | Asian | 35/67 | N/A | 32/61.54 |
| Yao Y, et al<sup>115</sup> | J Intensive Care | China | 63#<br>185# | 63.0 ± 13.4 | Asian | 135/54.4 | 0.35 (0.23, 0.42)*<br>1.69 (0.91, 5.06)* | 17/9.2 |
| Zhang JJ, et al<sup>46</sup> | Allergy | China | 140 | 57 (25, 87)* | Asian | 71/50.7 | 0.2 (0.1, 0.5)* | N/A |
| Zhang L, et al<sup>47</sup> | J Thromb Haemost | China | 343 | 62 (48, 69)* | Asian | 169/49.27 | 0.54 (0.20, 1.41)* | 13/3.8 |
| Zhang Y, et al<sup>2</sup> | NEJM | China | 3 | 68 (65 - 70) | Asian | 2/66.67 | 9.02 ± 10.37 | N/A |
| Zhou F, et al<sup>1</sup> | Lancet | China | 191 | 56 (46, 67)* | Asian | 119/62 | 0.8 (0.4, 3.2)* | 54/28.3 |
| Zou Y, et al<sup>48</sup> | BioSci Trend | China | 303 | 51 (16 - 88) | Asian | 158/52.15 | 0.45 (0.31, 0.81)* | N/A |

### Criteria for study selection and data extraction

All eligible studies meeting the following criteria were included: 1) the species was human, 2) the publications were original clinical investigations, and 3) the results were presented as or could be converted or digitized to mean ± SD (SDM) or percentage. Studies were excluded if they were: 1) reviews or editorial, single case reports, commentaries, or preclinical studies; 2) results that could not be converted or digitized to SDM or percentage, and 3) full articles or clinical data that were not available. Raw data of each sub-group at admission were extracted by ZLS and RZZ. Individual data of case series^2,32^, median and interquartile range^1,4-8, 13,33-47^, and ranges of median or mean^9,48,49^ were converted to SDM as described previously^50,51^. Percentages and SDMs were extracted directly from studies if available.

### Meta-analysis

To perform meta-analysis with the STATA v.16.1, the studies with two groups or more were pooled to compute weighted mean differences (WMD) and 95% confidence intervals (95% CI)^50^. Publication bias between selected studies was assessed with both the Egger’s and Begg’s tests using the metabias program. The stability of the results was confirmed by the random-effects trim and fill analysis using the metatrim program.

### Meta-regression analysis

The associations between D-dimer and demographic features, comorbidities, laboratory tests, radiographic results, treatments, hospitalization, outcomes, and complications were analyzed. D-dimer was considered as a covariate of other clinical variables. The standard errors of D-dimer were used to indicate the within-study variability, and the random-effect ReML method was applied. If observations were lesser than six, the results of this parameter were removed.

## Results

### 1. General information

Following the PRISMA guideline, we included 42 key studies for meta-analysis and meta-regression (**Figure S1**). The demographic features, D-dimer, ARDS, and mortality, were summarized in **Table 1**. In total, there were 14,862 laboratory-confirmed patients: 31 studies from China (5,961cases), 5 from USA (8,525 cases), 3 from Italy (62 cases), and 1 from Ireland (83 cases), France (150 cases), and Brazil (81 cases), respectively. The age was from 1 to 98-year-old. The races included Asian, African, Caucasian, and mixed with an incidence of ARDS ranged from 0 to 100%, and a fatality ranged from 0 to 95.3%. D-dimer level ranged from 0 to 35.7 μg/mL, and 9 studies reported a normal value (< 0.5 μg/mL).

### 2. Significant publication bias caused by divergent study designs

COVID-19, as a new emerging infectious disease, the included clinical studies had divergent designs. We compared the D-dimer level between relatively mild and severe groups of 23 studies (**Figure 1A**). Only a small increase of D-dimer, 0.97 μg/mL (95% CI 0.65, 1.29) was observed in the relatively severe group with significant publication bias (92.5%), which was corroborated by both the Begg’s (P=0.009) and Egger’s tests (P<0.001) (**Figure 1B**) and the random-effects filled funnel plot (**Figure 1C**, P<0.001). Similarly, significant variants caused by retrospective studies and case series were observed for both age (**Figure S2**) and mortality (**Figure S3**). Thus, it was considered to be inappropriate to perform meta-analysis without well-designed RCT (randomized controlled trials) studies. Instead, we hypothesized that D-dimer can serve as a critical covariate for clinical features.

**Figure 1.**
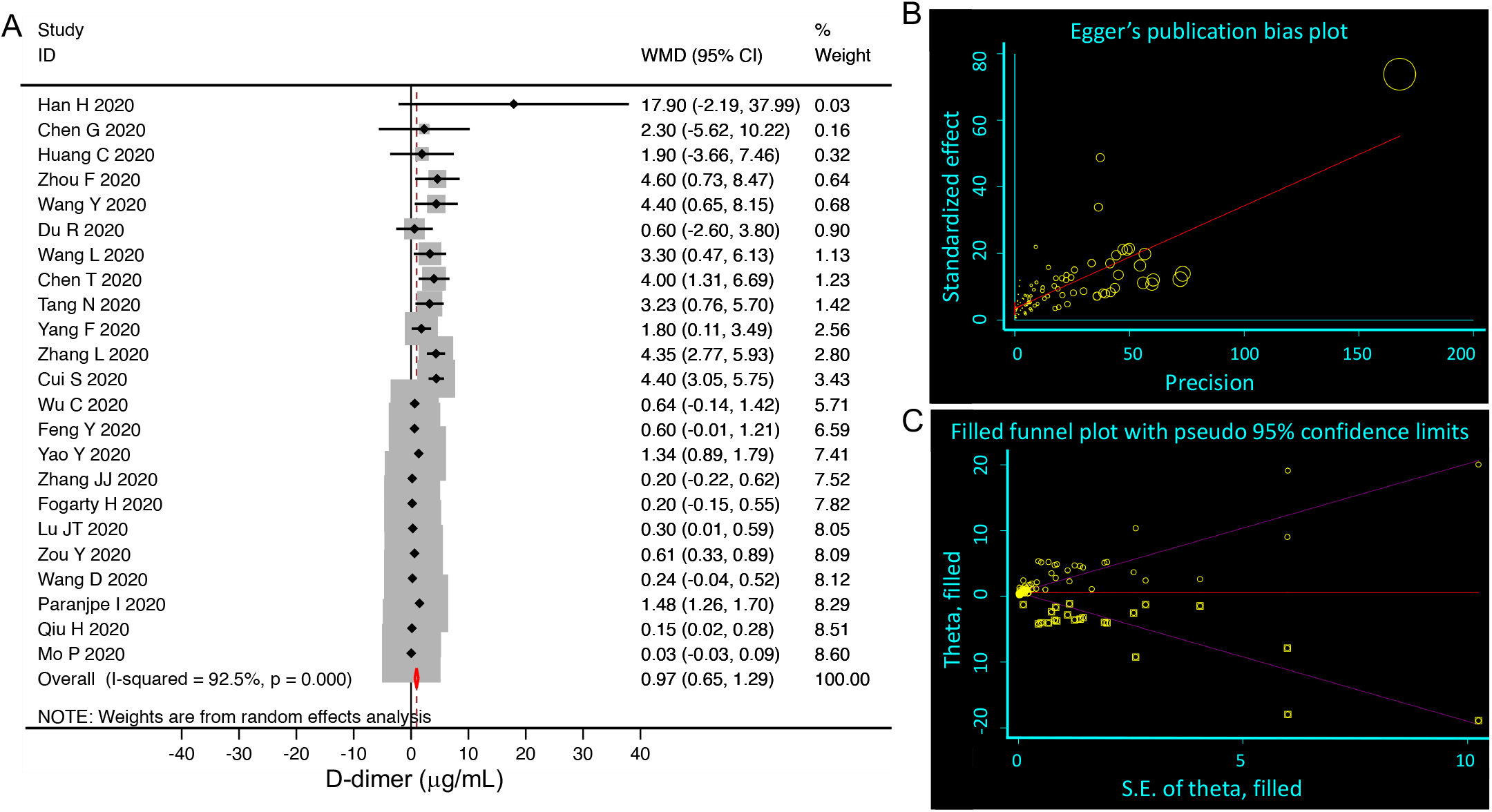
Random-effects meta-analysis of D-dimer level. **A**. Forest plot. We selected studies that had subgroups, which could be divided into relatively mild (including normal) and severe groups. We pooled weighted mean differences (WMD, black diamond, and gray square) and 95% CI (horizontal lines through the diamonds) of D-dimer from eligible 23 studies. Studies with only one group or moderate group were excluded. The red diamond represents the overall WMD. **B**. Egger’s publication bias plot. N=89, P<0.001. **C**. Filled funnel plot. P<0.001. Circle, raw data; square, pseudo data needed for symmetric distribution.

### 3. D-dimer correlated to demographic features

To detect the potential associations of D-dimer with 36 demographic characteristics of COVID-19 patients, we performed meta-regression (**Table S2**). Of these, preexisting medical conditions, including any comorbidity, hypertension, diabetes, chronic lung diseases, and cerebrovascular diseases, were positively associated with D-dimer (P<0.05). Age, gender, blood pressure, and dyspnea/tachypnoea positively correlated with D-dimer (P<0.05, **Table 2**). Moreover, the percentage of female and diastolic pressure was negatively correlated to D-dimer. Subgroup analysis showed a cutoff age was <65 years in the studies with two groups (<65 or ≥65), and further <50 in the studies with four groups (<50, ≥50, <60, ≥60, and ≥70) for a negative coefficient value (P<0.05, **Table S3**).

**Table 2.**
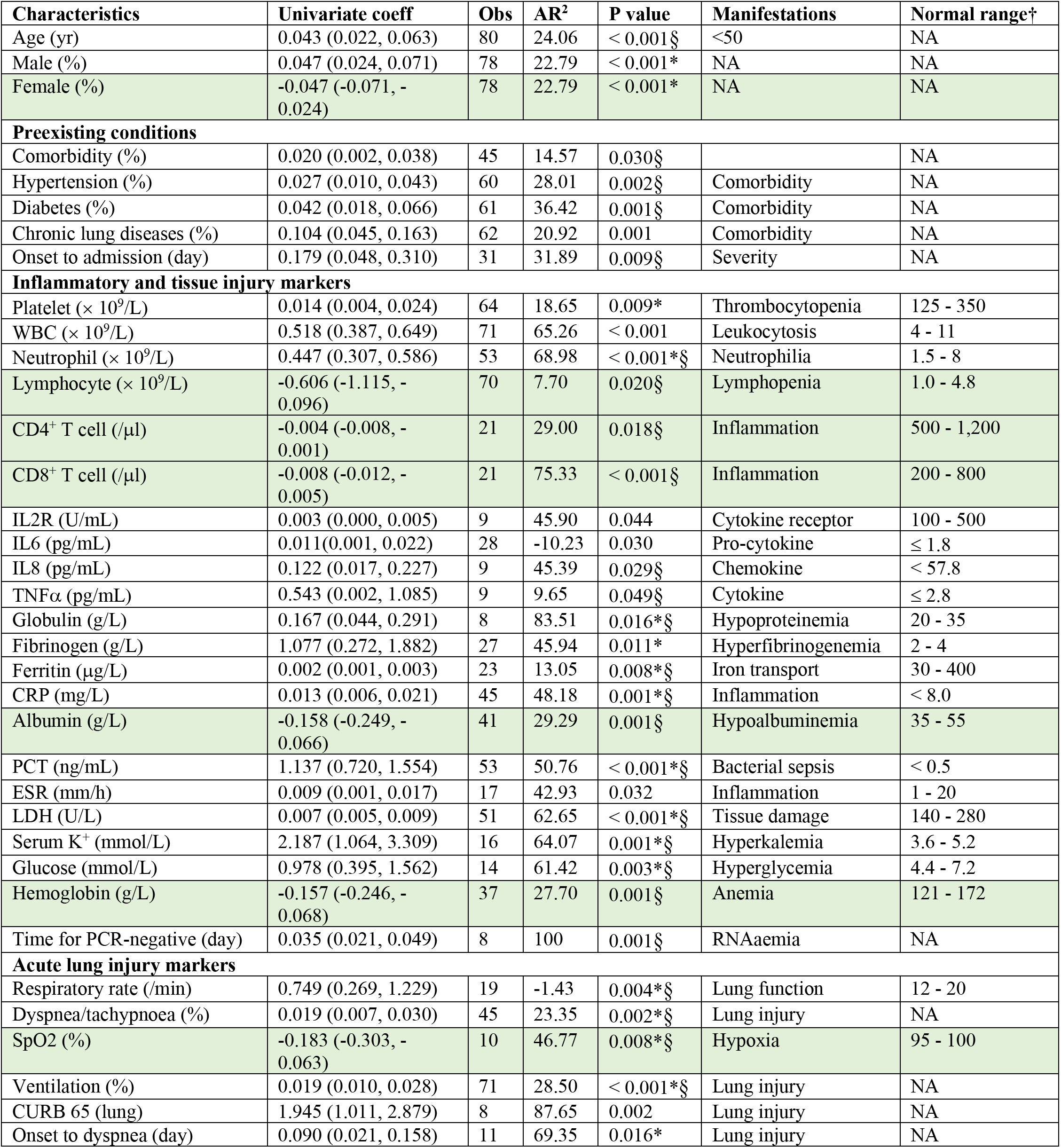
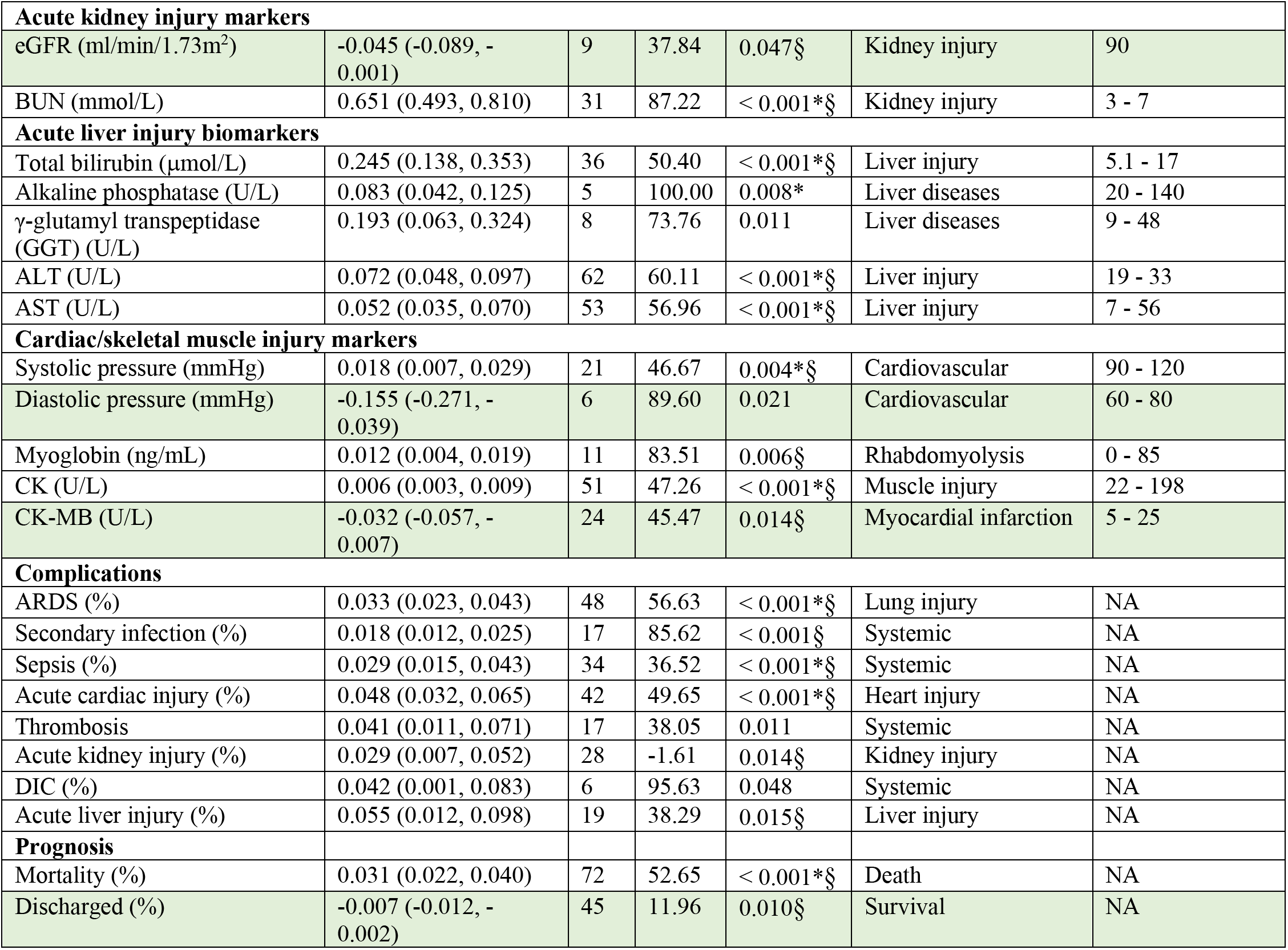
D-dimer associated clinical variables (58 of 106 in total with a sample size > 5, P < 0.05) identified by univariate-regression. Abbreviations: CI, confidence interval. Obs, the number of groups from the included studies. WBC, white blood cells. CK, creatine kinase. CK-MB, creatine kinase myocardial band. ALT, alanine aminotransferase. AST, aspartate aminotransferase. BUN, blood urine nitrogen. eGFR, estimated glomerular filtration rate. LDH, lactate dehydrogenase. PCT, Procalcitonin. CRP, C-reactive protein. ESR, erythrocyte sedimentation rate. IL2R, interleukin 2 receptor. TNFα, tissue necrosis factor a. CURB 65, CURB-65 Severity Score. ARDS, acute respiratory distress syndrome. TE, thromboembolism. DIC, Disseminated intravascular coagulation. † include the range of both male and female. P < 0.05 after removing the confounding variable age (*) or male (§) using bivariate regression.

### 4. D-dimer is a correlate of laboratory tests but not radiologic readouts

To analyze the correlation of D-dimer with 62 laboratory tests and radiological readouts, meta-regressions were conducted for individual variable and summarized in **Table S4**. There were 32 laboratory tests significantly associated with D-dimer (**Table S4**). In addition, FDP tended to associate with D-dimer levels (P=0.058, N=10). These tests could be roughly grouped as 1) inflammation and tissue injury markers, 2) acute lung injury markers, 3) acute kidney injury markers, 4) acute liver injury markers, and 5) cardiac/skeletal muscle injury markers (**Table 2**).

### 5. D-dimer is associated with mechanical ventilation

To evaluate the relationship between interventions and D-dimer, 11 therapies were studied in association with the D-dimer level (**Tables S5 and 2**). Interestingly, mechanical ventilation was positively associated with D-dimer (N=71, P<0.001, **Figure 2**). Immune enhancement therapy (P=0.084, N=24) was also found to correlate with D-dimer if additional observations were available to increase sample size.

**Figure 2.**
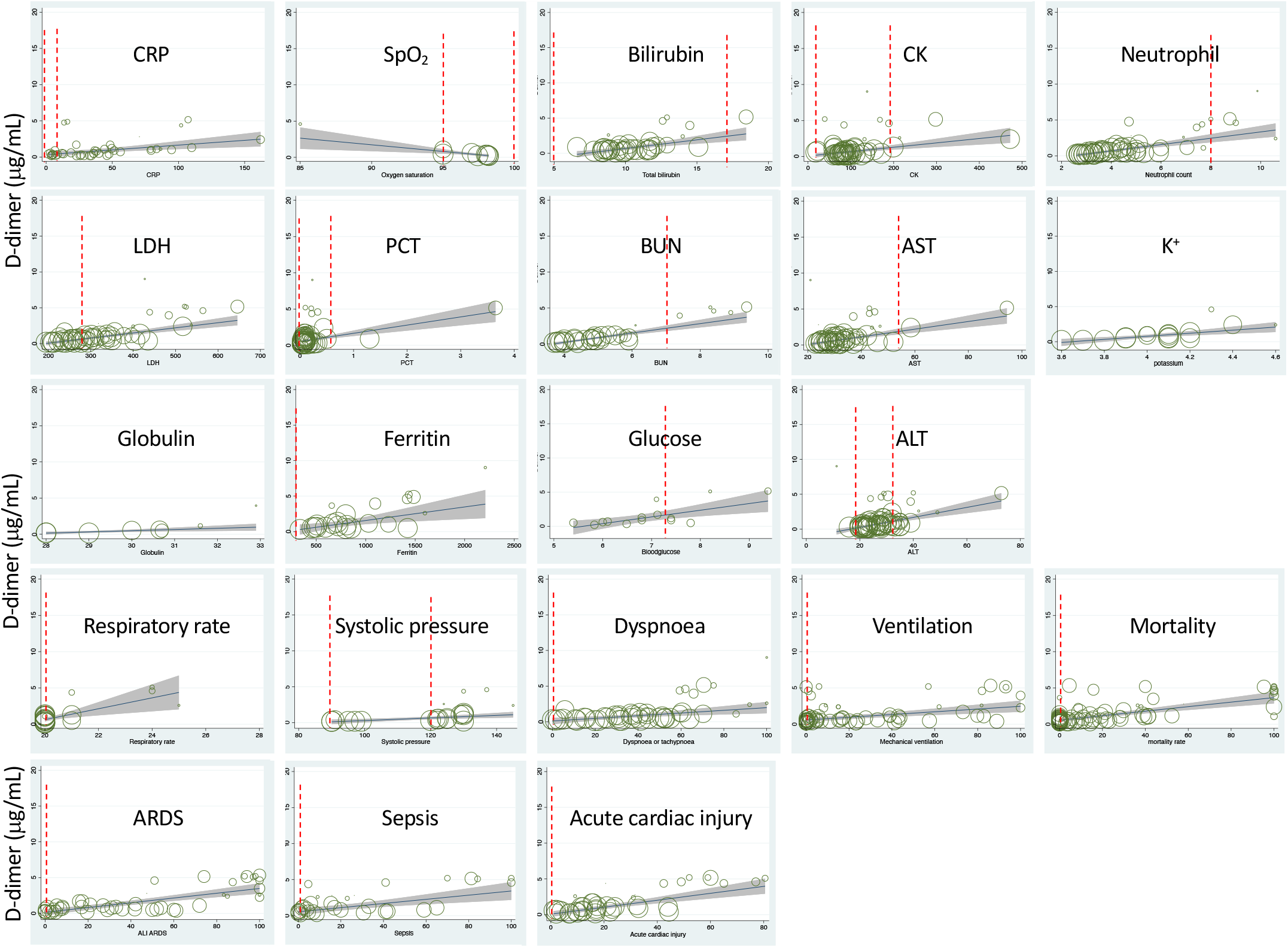
Random-effects meta-regression of D-dimer associated variables.

### 6. D-dimer serves as an independent risk factor for fatal organ injury and systemic conditions

To examine if D-dimer is an independent risk factor for deadly complications, the dependence of fatal organ injury and systemic disorders was analyzed using the metareg program. The results were summarized in **Tables S6** and **2**. Acute lung (ALI/ARDS), heart, kidney, and liver injuries were significantly associated with D-dimer (P<0.05, **Figure 2**). In addition, four systemic complications, i.e., sepsis, secondary infection, disseminated intravascular coagulation (DIC), and coagulopathy showed marked associations with D-dimer. Acute brain injury and acidosis showed a tendency to associate with the D-dimer. Together, both acute fatal organ injury and systemic complications could be predicted by D-dimer.

### 7. D-dimer predicts severity, hospitalization, and outcomes

Given the correlation of D-dimer with the demographic features, abnormal laboratory tests, interventions, and severe complications, we hypothesized that D-dimer is an independent indicator for these disease progression (CURB 65 score, onset to admission, onset to dyspnea), hospitalization (discharged, time taken to turn SARS-CoV-2 PCR negative), and mortality. We analyzed the correlation of D-dimer and these clinical readouts and summarized in **Tables S7** and **2**. D-dimer was positively associated with the severity of lung injury (CURB 65), the days from the onset to admission, onset to dyspnea, time taken to be PCR negative, and overall mortality (**Figure 2**). In contrast, the discharge rate was negatively related to D-dimer, demonstrating the capability of D-dimer to serve as a prognostic variate for outcomes.

### 8. Exclusion of age and gender as confounding factors

Age and gender have been identified as preexisting medical conditions associated with COVID-19 resulting in higher mortality^3,34,47^. We eliminated their effects on the association of D-dimer with 61 identified variables in bivariate meta-regression analyses. Age was a significant confounding factor of 27 variables, and 26 variables were still associated with the D-dimer independent on age (**Table S8**). In contrast, 31 variables were disassociated with the D-dimer. By comparison with age, male gender was a much weaker confounding covariate (**Table S9**). Males were significantly associated with 11 variables and results in the dissociation of 14 variables with D-dimer. The association of 28 and 42 variables with D-dimer was still significant after considering age and male as a covariate, respectively (**Table 2**). Age-affected 18 variables include comorbidity, hypertension, diabetes, lymphocyte, CD4^+^ and CD8^+^ T cells, IL8, TNFα, eGFR, hemoglobin, albumin, CK-MB, onset to admission, onset to PCR negative, discharged, secondary infection, acute kidney injury, and acute liver injury. There were 10 variables confounded by both age and male, including diastolic pressure, chronic lung diseases, WBC, IL2R, IL6, ESR, GGT, CURB 65 score, coagulopathy, and DIC. These four variables were markedly affected by covariate male: platelets, fibrinogen, alkaline phosphatase, and onset to dyspnea. Finally, 22 variables were significantly associated with D-dimer: respiratory rate, systolic pressure, dyspnea, serum K^+^, neutrophils, globulin, CRP, ferritin, LDH, PCT, SpO_2_, blood glucose, BUN, total bilirubin, ALT, AST, Ck, mortality, ventilation, ARDS, sepsis, and acute cardiac injury (**Figure 3**).

**Figure 3.**
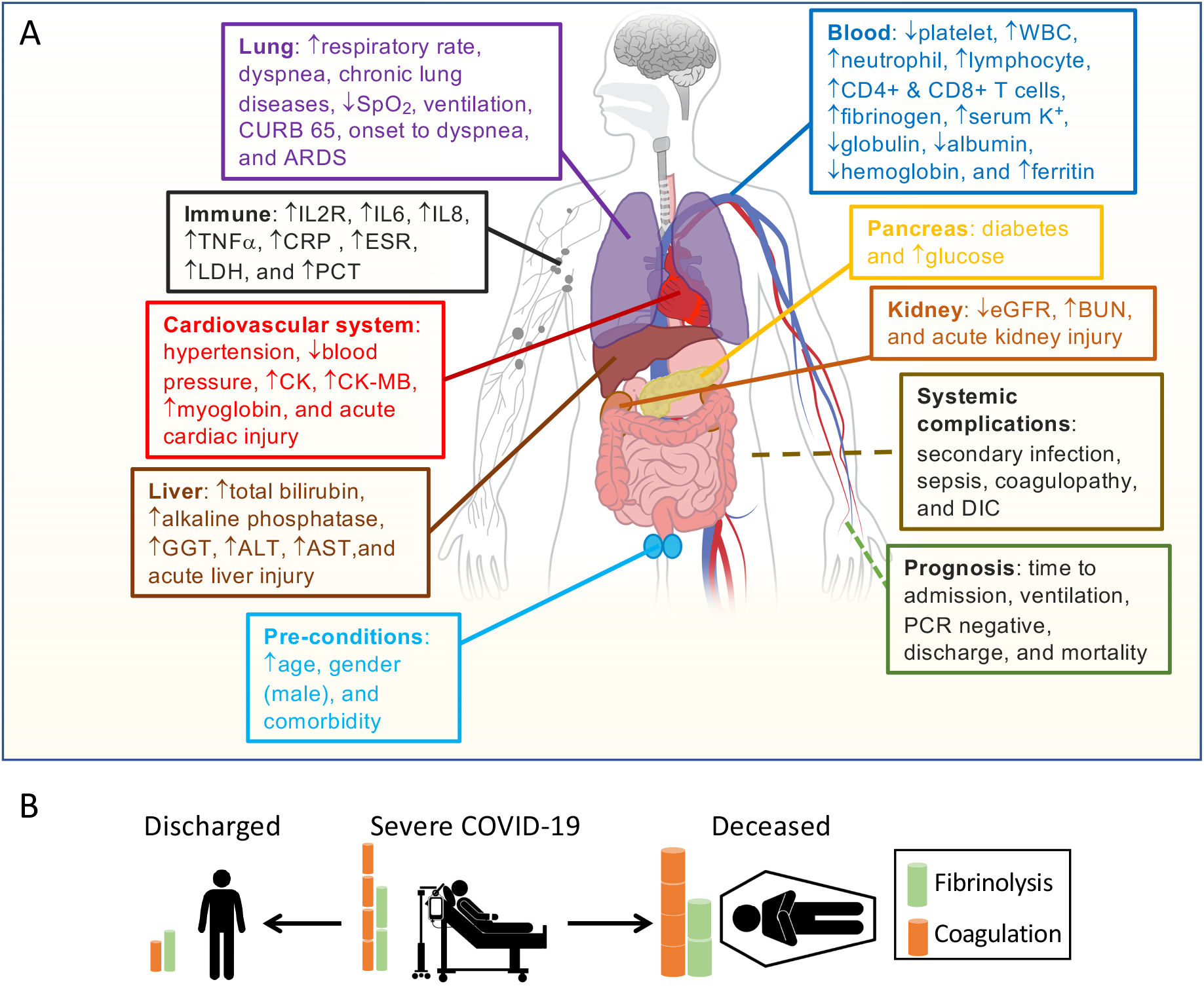
Schematic summary of D-dimer related variables. **A**. Demography, radiology, laboratory tests, complications, and outcomes sorted by organ functions. **B**. Hemostasis and outcomes of COVID-19 patients.

## Discussion

We aimed to systematically analyze the relationships between circulating D-dimer level in the blood on admission and clinical variables to test for the underlying mechanisms for the dysfunctional fibrinolysis in critically ill COVID-19 patients. There is a range of plasma D-dimer levels on hospital admission. The directions of dynamically changed FDPs for hospitalized patients are different between discharged and deceased cohorts. As revealed by our meta-regression analysis, plasma D-dimer is associated with comorbidities, demographics, some laboratory tests, radiology, hospitalization, complications, and outcomes. These results suggest that in addition to serving as an independent predictor for fatality, severity, and could potentially serve as a marker for daily monitoring thrombolytic therapy, D-dimer is a non-specific biomarker that interacts with other coagulation molecules, inflammatory cytokines, and markers for organ/tissue injury. Of note, the interplay of acute-phase proteins with fibrinogen and D-dimer suggests that infection-induced inflammation (cytokines and chemokines) initiates a state of hyperfibrinolysis. This notion is supported by the disassociation of D-dimer with the entire coagulation panel (PT, APTT, factor VIII and XI, TAT)^52,53^. In general, hyperfibrinolytic homeostasis maintains vascular patency and normal organ function under physiological conditions (**Figure 3B**). SARS-CoV-2 and co-bacterial infection initiates a hypercoagulable state followed by hyperfibrinolysis in COVID-19. If hyperfibrinolysis can counter excessive coagulopathy, then the patients could be protected against thrombosis. Otherwise, insufficient local hyperfibrinolysis in the lung of non-survivors will be exhausted.

### 1. Clinicopathological mechanisms of fibrinolysis in COVID-19

Soluble fibrinogen is synthesized in the liver (1.7-5g/d) primarily and others, including the bone marrow, brain, lung, and gastrointestinal epithelium (**Figure 4A**). It mainly distributes in the plasma (75%), interstitial fluid (16%), platelets, and lymph^54,55^. IL6 and other proinflammatory cytokines/chemokines, steroids, and miRNAs upregulate the fibrinogen synthesis up to 10-fold during the acute phase of injury and infection. Fibrinogen (2-3%) can be turned over to fibrin monomers by thrombin and cleaved by plasmin, the process termed fibrinogenolysis^54,56^. Fibrinogen degradation products (FgDP) are 2- or 3-fold that of plasma D-dimer but with a shorter half lifetime (2.8 h vs 16 h for D-dimer)^54,56^. Crosslinked fibrin is formed in the presence of FXIIIa. At the endothelial cell surface of injured blood vessels, fibrin(ogen) “glues” the plugs formed by the aggregation of platelets to develop thrombi (clots). Excessive fibrin deposition and inflammation activate endothelial cells to produce tPA and urokinase plasminogen activator (uPA)^54,55^. Either tPA or uPA is capable of cleaving hepatocyte-derived plasminogen to activate plasmin. Plasmin proteolytically cleaves fibrin within the thrombi into FDP and D-dimers, the end products of fibrinolysis. Given the very short half lifetime (in seconds or minutes) of endogenous thrombin, tPA, uPA, and plasmin, and the overwhelming antithrombin, plasminogen activator inhibitor 1 (PAI-1), and plasmin inhibitors with a much longer lifetime in the plasma^57^, the primary cleavage of plasminogen and fibrin may predominately take place at the surface of clots. Eventually, FgDP and FDP (D-dimer) will be catabolized in the liver and captured by the reticuloendothelial system and excreted to the bile^54,55^. Another clearance pathway is via the kidney to excreted to urine^54,55^. D-dimer assay has routinely applied for excluding PE, deep venous thromboembolism, and DIC, as well as a marker for monitoring the effects of fibrinolytic/thrombolytic therapy.

**Figure 4.**
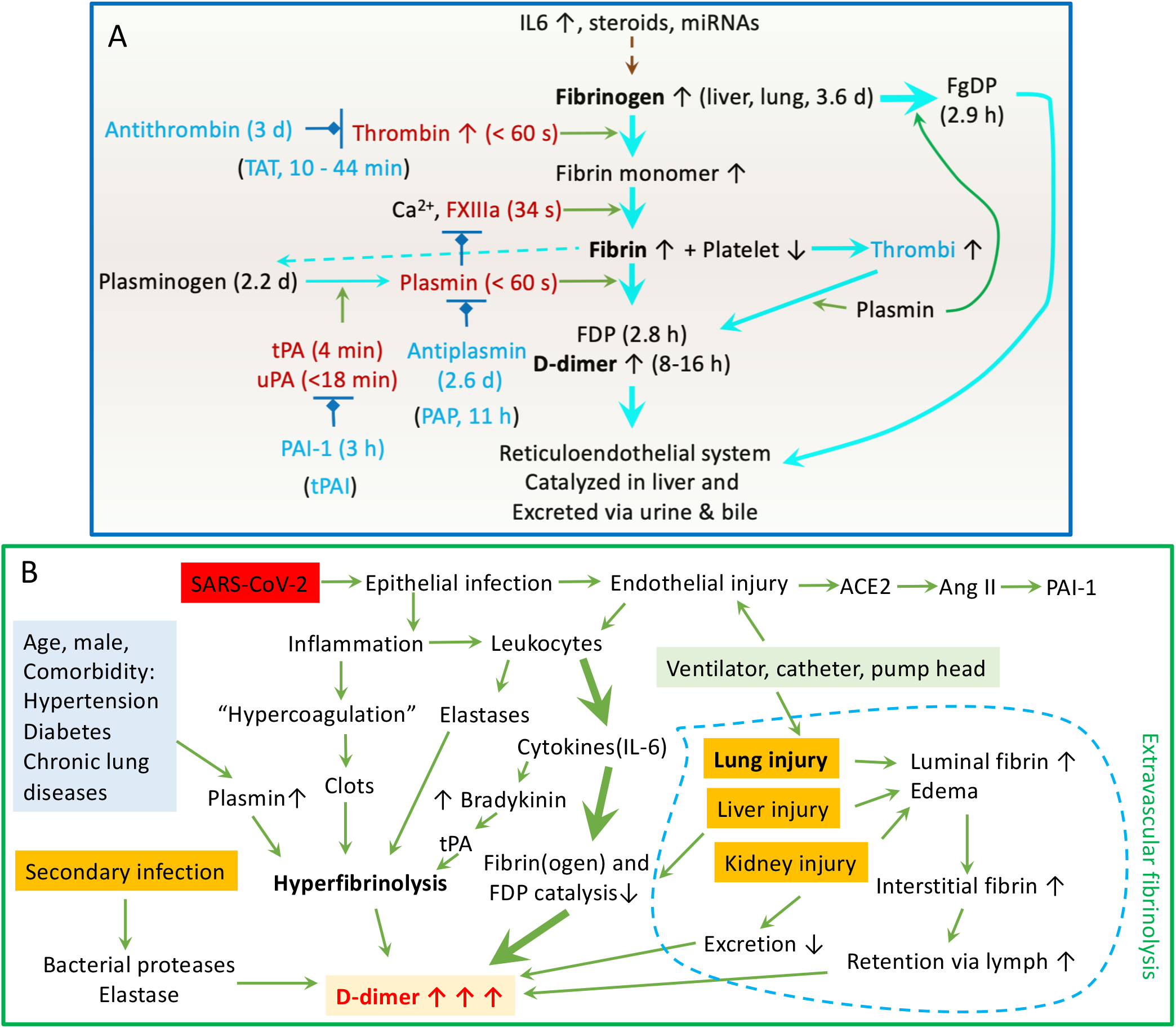
Clinicopathological mechanisms of hyperfibrinolysis in COVID-19. **A**. Fibrinolysis system with half lifetime for key components. **B**. Potential mechanisms for elevated D-dimer in COVID-19.

Age is associated with an increased D-dimer level in COVID-19 patients at admission as a covariate or independent prognostic marker for the outcomes of COVID-19. The cutoff value of D-dimer (0.5 μg/mL) is age-dependent for healthy cohorts^58,59^. Our subgroup analysis is consistent with the concept that adults older than 50 approach the threshold of the D-dimer levels seen in normalcy (**Table S9**). The difference in D-dimer between men and women is minor in healthy population^59^. The positive association of D-dimer with the percentage of male patients in COVID studies suggests more severe cases in men than women when admitted.

#### Mechanisms for increased baseline D-dimer at admission

We demonstrate that patients with comorbidities, including overall comorbidities, hypertension, diabetes, and chronic respiratory diseases, correlates with D-dimer levels at admission. The association can be explained by the comorbidity-associated elevations in plasmin^60^. Increased D-dimer has been reported in the following conditions^56,59,61^: DIC, VTE, pregnancy, advance age, malignancy, sepsis, burns, trauma, myocardial injury, cerebrovascular event, liver disease, severe renal disease, surgery, ARDS, pancreatitis, diabetes, Alzheimer, disability, and the neonatal period. The different time points for admission after disease onset and D-dimer testing could partially account for the different D-dimer levels between COVID-19 groups and the studies. This is in fact the rationale to apply meta-regression analyses. Patients with severe COVID-19 may be those who were ill for a longer period than mild controls. This is supported by the association of the D-dimer level and the days from onset to admission (**Table 2**). Alternatively, the progression of disease may be much quicker in critically ill patients at admission. A significant increase in plasma tPA was reported in ICU patients with elevated D-dimer^62,63^. In contrast, the a2-antiplasmin protein was at the upper limit of normal level^62^.

#### Synthesis, metabolism, distribution, and excretion offibrin(ogen)

Our regression analysis reveals that elevated D-dimer is associated with a broad spectrum of immune responses to SARS-CoV-2 infection, including increased pro-inflammatory cytokines (IL2R, IL6, IL8, and TNFα), acute phase proteins (CRP/C-reactive protein, fibrinogen, ferritin, and albumin), and inflammation indicators (ESR-erythrocyte sedimentation rate, PCT-procalcitonin, globulin, white blood cells, neutrophil, lymphocyte, and CD4^+^ & CD8^+^ T cells). Moreover, the days for reversion of the PCR test to negative is related to the D-dimer level. These correlations support the concept that interactions between high levels of circulating cytokines and hyperfibrinolysis may be functionally correlated. The binding of the spike proteins of SARS-CoV-2 to the ACE2 receptor in host respiratory epithelial cells downregulates the protective ACE2/Ang1-7/Mas axis, leading to increased expression of PAI-1^64^. Airway and lung epitheliitis releases proinflammatory cytokines to attract leukocytes from the blood. Moreover, infiltrated immune cells are activated and unleashed to attach normal lung tissues by releasing overwhelming cytokines. These cytokines upregulate positive acute-phase protein (i.e., fibrinogen), TF, and trypsin expression and inhibit negative proteins (i.e., albumin). Trypsin-activated matrix metalloproteinases break down the basolateral membrane and interstitial extracellular matrix. Further, endotheliopathy occurs in infected capillaries to initiate a local hypercoagulable state. The kinin-bradykinin is activated by IL6 to stimulate tPA expression in endothelial cells^64^. Deposition of fibrin (clotting) activates endothelial cells to express more IL8, which suppresses clot lysis time^65^. This, combined with the hypoxia-causing eryptosis, may be the reason for IL8 associating with high mortality. ESR is associated with the severity of COVID-19 patients^66^. In addition to carrying oxygen, erythrocyte-bound streptokinase and tPA break down the clots. Extrathyroidal produced PCT if maintained at an elevated level by cytokines is an indicator of poor outcomes of COVID-19^67^. It has been used as a marker for co-bacterial infection in septic shock and influenza. Neutrophil extracellular traps (NETs) may contribute to organ damage and promote thrombosis and fibrinolysis via elastase, so do lymphocytes/macrophages. Hepatocyte-synthesized globulins and albumin are involved in liver function, coagulation, and antiinflammation. Their reduced levels by vast consumption predict a poor outcome. Albumin acts as anticoagulant and antiplatelets and increases vascular permeability^68^, which seems to explain the link between elevated D-dimer and hypoalbuminemia (**Figure 4B**).

#### Coagulation and fibrinolysis

Our results show a close relation between the plasma D-dimer and thrombocytopenia, hyperfibrinogenemia, and thrombosis. Significant reductions in platelets could be due to 1) vast consumption for adhesion and aggregation to the surface of injured lung capillaries, 2) a decrease in platelet release from fully mature megakaryocytes in the injured lung, and 3) abnormal hematopoiesis of infected bone marrow. Hypercoagulation and abnormal fibrinolysis are inconsistent in COVID-19 patients. There is no correlation between abnormal coagulation variables and thrombosis except elevated D-dimer^53^. The hypercoagulable state is systemic, but the hyperfibrinolysis occurs in the pulmonary capillaries. This is supported by the “fibrinolysis resistance” or “fibrinolysis shutdown” in vitro whole blood clot lysis^22,69,70^. The moderate reduction in circulating plasminogen of ICU COVID-19 patients eliminates the thrombolytic effects of tPA^71^, which is supported by thromboelastometry (TEM)^49,72^. As the half-life of plasminogen is 2.2 days, the fibrinolysis resistance in vitro cannot be explained by the distance to the clots in vivo (about 45 seconds per cycle through the circulation system) and the time between collection of blood to assay. Indeed, sequential TEM tests on days 0, 5, and 10 found no fibrinolytic activity too^72^, probably due to extremely elevated PAI-1 and insufficient supply of tPA. Another line of evidence is the dynamic changes in increased TAT (thrombin-antithrombin) and reduced PAP (plasmin-antiplasmin) complexes in non-survivors^73^, a sign of hyperactive plasminogen cleavage and resultant hyperfibrinolysis. This is also probably due to the adhesion of plasminogen to the clot at the cell surface and/or impaired circulation through the pulmonary capillaries blocked by microthrombi. Thrombin level was significantly reduced in COVID-19 patients associated with increased D-dimer^62^. Of note, spontaneous fibrinolysis was enhanced by high tPA plasma of a subset COVID-19 patients^63^.

#### Cell/tissue injury

Lysis of platelets, erythrocytes, and other cells caused by infection contribute to higher serum K^+^ levels and reduced hemoglobin. The tissue damage is supported by the increased LDH level, together with serum K^+^ and hemoglobin correlate with D-dimer. A decrease in renal excretion of K^+^ salts may worsen the hyperkalemia in COVID-19. Cell death at the clotting site and alveoli increase the release of fibrin(ogen) into the blood and the risk of embolism. Meanwhile, a new reaction surface is exposed for producing D-dimer by plasmin both locally and systemically.

#### Acute lung injury

D-dimer is associated with acute lung injury/ARDS, including hypoxia (respiratory rate, dyspnea, SpO_2_, CURB 65, and the onset of dyspnea) and the need for mechanical ventilatory support. The lung is a key early target organ of SARS-CoV-2 infection and could be a crucial source of D-dimer for some COVID-19 patients or at some timepoints. At the early stage before vascular endothelialitis, extravascular fibrin degradation in the alveolar sacs and interstitium could be the major site responsible for the generation of elevated D-dimer levels. uPA in the exudate cleaves plasminogen to generate plasmin, together with elastases released from macrophage and neutrophils to resolve fibrin, but the process is greatly inhibited by locally increased levels of PAI-1 in most forms of ARDS ^74-76^ and likely in that induced by COVID-19. Both fibrin and FDPs will be retained to the venous system via the lymphatic system (17L/day). Lymphatic retention may take place in other edematous organs until the patients are either recovered or expired^77,78^. This may be the case for patients with elevated D-dimer but not vascular thromboembolism. Disassociations of increased plasma D-dimer with either augmented tPA or PAI-1 in relatively young COVID-19 patients (age 41.7 ± 14) supports the importance of extravascular fibrinolysis^63^. This is supported by the benefits of inhaled fibrinolytics^79-81^, postmortem findings^15-19^, and clinical features. Vasculopathy in the pulmonary capillaries further enhances the local hyperfibrinolytic process. Plasma D-dimer and other FDPs rise along with the pulmonary hypercoagulable state and a proinflammatory state. Hypoxia occurs in patients with the lungs with diffuse alveolar damage, a result in part from fibrin deposition and degradation. Mechanical ventilation with positive pressure could facilitates the retention of extravascular generated D-dimers.

#### Acute liver injury

The results show an association of acute liver injury and elevated D-dimer in the blood. Postmortem biopsies have detected congestion, vascular changes, and periportal lymphocyte infiltration and fibrosis^15,17,18^. The D-dimer level is increased in various liver diseases, as the result of the overwhelming deposition of fibrin(ogen) and subsequent inflammation injury in the liver, which leading to suppressed catabolism of D-dimer.

#### Acute kidney injury

The association of D-dimer and estimated glomerular rate (eGFR) in COVID-19 patients is supported by the clinical practice for adjusting D-dimer cutoff value for patients with renal dysfunction^82^. Reduced eGFR will reduce the renal clearance of D-dimer and result in an elevated plasma D-dimer level^83^. On the other hand, increased blood urea nitrogen (BUN) level (cutoff: >4.6 mmol/L) as well as D-dimer (cutoff: ≥0.845 μg/mL) serve as independent risk factors for predicting in-hospital mortality^84^. Coronavirus-like particles were detected in damaged kidneys^19,85^ and may have stimulated renal uPA synthesis.

#### Acute cardiac injury

The heart can be a significant target of SARS-CoV-2 infection in some patients, and ACE2 receptors are expressed in myocytes. Some patients showed lymphocytic myocarditis and myositis^19,86^. Approximately 20% of postmortem samples from the heart had fibrin thrombi and interstitial edema^86^. Thus, hypoxic myocardial diseases, including heart stroke, myocardial infarction caused by infection, resulting in an incremental increase in plasma D-dimer^87^. The acute cardiac injury was detected by elevated CK, CK-MB, and myoglobin. Of note, a 16-year LIPID trial demonstrates that higher D-dimer is an independent predictor of all-cause mortality, cardiovascular mortality, cancer mortality, and other fatal events^88^.

#### Hyperglycemia

Nearly half of COVID-19 patients (47%) had elevated levels of blood glucose at admission^89^. Hyperglycemia is an independent risk factor for progression to critical cases and in-hospital deaths for both mild and severe patients. Intriguingly, stress hyperglycemia is an independent risk factor associated with worse outcomes among critically ill patients^90^. SARS-CoV-2 induced cytokine storm, and the direct infection of the pancreas leads to insulin resistance. Hyperglycemia then results in increased TF, FVII, thrombin and PAI-1 levels, promoting clot formation and restraining fibrinolysis. Glycation and oxidation of these coagulation and fibrinolysis proteins in compact clots develop resistance to plasmin^91^.

#### Systemic complications

Systemic complications, including bacterial co-infection, septic shock, DIC, and thromboembolic events in large blood vessels, have a high incidences and poor outcomes in non-survivors. D-dimer is associated with these severe complications in critically ill patients. All of the mechanisms illustrated in **Figure 4B** appear to be involved in hyperfibrinolysis at the late stage of disease.

#### Sudden increments in the D-dimer level in ICU patients

A rapid rise in D-dimer level was described in non-survivors, particularly for the last test before death. It can be caused by the rapid exacerbation of septic organ failures, DIC, and secondary infection. Pump head thrombosis in patients treated with ECMO^28^, ventilation^14,92^, steroids, tocilizumab, and tPA administration leads to a transient augmentation in plasma D-dimer^12^.

### 2. Prognostic relevance

Our results showed a strong positive association of elevated D-dimer on admission with mortality, indicating the prognostic value of an elevated D-dimer for the high risk of death. This is further corroborated by the positive correlation between D-dimer and days from onset to admission, the need for ventilation, and the days taken for PCR test reversion to negative. Another line of supportive evidence is the negative relation of D-dimer and discharge probability. This data shows that prompt admission and clearance of SARS-CoV-2 virus may alleviate the severity and reduce fatal events by preventing the hyperfibrinolysis and inflammation. D-dimer, as a prognostic marker, is supported by other reviews^93-96^ and clinical studies^1,47^. However, given the dynamic feature of plasma D-dimer, the peak D-dimer value improves the prognostic relevance^97-101^. Combination with other clinical parameters of biomarkers, for example, IL6 or chest imaging, may be of value for the diagnosis and follow-up of PE and VTE because D-dimer could also be increased by extravascular fibrinolysis^77^.

### 3. Strengths

We have systematically reviewed the literature using meta-regressions and meta-analyses to define clinicopathologic mechanisms of fibrinolysis with elevated D-dimer and its diagnostic and prognostic relevance. Based on the findings, we infer that this test may be tractable for the stratification and interventions of COVID-19 patients. The associations of D-dimer and other clinical variables indicate either a relationship that may be cause and effect, or indirect.

Some of the D-dimer-associated variables have been confirmed or could be prognostic biomarkers for developing fatal events and in-hospital mortality. Extremely elevated plasma D-dimer seems to be the consequence of hyperfibrinolysis predominately in the pulmonary capillaries and other organs. A dynamic increase in the D-dimer level may be associated with thromboembolism and higher fatality, while we infer that a continuous decline by daily testing will generally lead to recovery.

### 4. Limitations

Because COVID-19 is a new emerging disease, most of the included studies are descriptive, case series, retrospective, single-center, and observational. Most of the studies included are from China/Asia, where the pandemic was discovered. Diversity in cohorts exists from children to seniors associated with variant comorbidities and complications. Inconsistent grouping strategies between studies pose a challenge for meta-analysis, for example, ICU vs non-ICU, ARDS vs non-ARDS, control vs COVID-19, survivor vs non-survivor, severe vs non-severe, VTE vs non-VTE, death vs recovered, mild vs moderate, moderate vs severe, normal vs abnormal D-dimer, etc. We cannot exclude the potential pre- and post-test deviations regarding the methodology for D-dimer assays. Meta-regression but not meta-analysis may partially mitigate these deviations.

### 5. Conclusions

We have reviewed the fibrinolytic mechanisms for the most common biomarker D-dimer in severe COVID-19 patients and propose a new model of insufficient pulmonary hyperfibrinolysis in both extra- and intravascular compartments. SARS-CoV-2 infection-initiated primary hypercoagulation occurs with hyperfibrinolysis in the lung and other end-organs, presenting with D-dimer elevations. Suppression of the excretion and catabolism in the injured liver and kidney likely contributes to the accumulation of D-dimer in the blood. This process is further enhanced by systemic septic DIC, multiorgan failure, and secondary infection in deceased patients. Patients who have survived show restoration of systemic hyperfibrinolysis over the hypercoagulable state, as indicated by normalized plasma D-dimer. Our model provides new information on the clinical relationship between aberrant fibrinolysis and D-dimers in patients with COVID-19.

## Data Availability

All data refered to in the manuscript and note links below are available.

## Contributors

HLJ, ZLS, and RZZ were responsible for searching the literature, reviewing extracted data, meta-analysing data, and preparing drafts of this manuscript. HLJ, SI, AK, SLL, MAM, and GHY improved the final manuscript, and all authors approved submission.

## Acknowledgments

This study was supported by NIH grants HL87017, HL095435, HL134828, HL154103, HL130402, AI112381, AI150473, and HL14285301.

## Reference

1. Zhou F, Yu T, Du R, et al. Clinical course and risk factors for mortality of adult inpatients with COVID-19 in Wuhan, China: a retrospective cohort study. Lancet 2020; 395(10229): 1054–62.

2. Zhang Y, Xiao M, Zhang S, et al. Coagulopathy and antiphospholipid antibodies in patients with COVID-19. N Engl J Med 2020; 382(17): e38.

3. Xie J, Hungerford D, Chen H, et al. Development and external validation of a prognostic multivariable model on admission for hospitalized patients with COVID-19. medRxiv 2020: 2020.03.28.20045997.

4. Xu XW, Wu XX, Jiang XG, et al. Clinical findings in a group of patients infected with the 2019 novel coronavirus (SARS-Cov-2) outside of Wuhan, China: retrospective case series. BMJ 2020; 368: m606.

5. Wang Y, Zhang D, Du G, et al. Remdesivir in adults with severe COVID-19: a randomised, double-blind, placebo-controlled, multicentre trial. Lancet 2020; 395(10236): 1569–78.

6. Wang D, Hu B, Hu C, et al. Clinical characteristics of 138 hospitalized patients with 2019 novel coronavirus-infected pneumonia in Wuhan, China. JAMA 2020; 323(11): 1061–9.

7. Richardson S, Hirsch JS, Narasimhan M, et al. Presenting characteristics, comorbidities, and outcomes among 5700 patients hospitalized with COVID-19 in the New York city area. JAMA 2020; 323(20): 2052–9.

8. Lu J, Hu S, Fan R, et al. ACP risk grade: a simple mortality index for patients with confirmed or suspected severe acute respiratory syndrome coronavirus 2 disease (COVID-19) during the early stage of outbreak in Wuhan, China. medRxiv 2020: 2020.02.20.20025510.

9. Akalin E, Azzi Y, Bartash R, et al. COVID-19 and kidney transplantation. NEngl J Med 2020; 382(25): 2475–7.

10. Bona RD, Valbusa A, Malfa G, et al. Systemic fibrinolysis for acute pulmonary embolism complicating acute respiratory distress syndrome in severe COVID-19: a case series. Eur Heart J Cardiovasc Pharmacother 2020: 10.1093/ehjcvp/pvaa087.

11. Ly A, Alessandri C, Skripkina E, et al. Rescue fibrinolysis in suspected massive pulmonary embolism during SARS-CoV-2 pandemic. Resuscitation 2020; 152: 86–8.

12. Chan KH, Slim J, Shaaban HS. Pulmonary embolism and increased levels of D-dimer in patients with coronavirus disease. Emerg Infect Dis 2020; 26(8): 1941–3.

13. Helms J, Tacquard C, Severac F, et al. High risk of thrombosis in patients with severe SARS-CoV-2 infection: a multicenter prospective cohort study. Intensive Care Med 2020; 46(6): 1089–98.

14. Connors JM, Levy JH. COVID-19 and its implications for thrombosis and anticoagulation. Blood 2020; 135(23): 2033–40.

15. Tian S, Xiong Y, Liu H, et al. Pathological study of the 2019 novel coronavirus disease (COVID-19) through postmortem core biopsies. Mod Pathol 2020; 33(6): 1007–14.

16. Dolhnikoff M, Duarte-Neto AN, de Almeida Monteiro RA, et al. Pathological evidence of pulmonary thrombotic phenomena in severe COVID-19. J Thromb Haemost 2020; 18(6): 1517–9.

17. Schaller T, Hirschbuhl K, Burkhardt K, et al. Postmortem examination of patients with COVID-19. JAMA 2020; 323(24): 2518–20.

18. Beigmohammadi MT, Jahanbin B, Safaei M, et al. Pathological findings of postmortem biopsies from lung, heart, and liver of 7 deceased COVID-19 patients. Int J Surg Pathol 2020: 10.1177/1066896920935195.

19. Bradley BT, Maioli H, Johnston R, et al. Histopathology and ultrastructural findings of fatal COVID-19 infections in Washington State: a case series. Lancet 2020; 396(10247): 320–32.

20. Al-Ani F, Chehade S, Lazo-Langner A. Thrombosis risk associated with COVID-19 infection. A scoping review. Thromb Res 2020; 192: 152–60.

21. Marchandot B, Sattler L, Jesel L, et al. COVID-19 related coagulopathy: a distinct entity? J Clin Med 2020; 9(6): 1651.

22. Wright FL, Vogler TO, Moore EE, et al. Fibrinolysis shutdown correlation with thromboembolic events in severe COVID-19 infection. J Am Coll Surg 2020; 231(2): 193–203.e1.

23. Wang J, Hajizadeh N, Moore EE, et al. Tissue plasminogen activator (tPA) treatment for COVID-19 associated acute respiratory distress syndrome (ARDS): A case series. J Thromb Haemost 2020; 18(7): 1752–5.

24. Barrett CD, Oren-Grinberg A, Chao E, et al. Rescue therapy for severe COVID-19 associated acute respiratory distress syndrome (ARDS) with tissue plasminogen activator (tPA): a case series. J Trauma Acute Care Surg 2020; 89(3): 453–7.

25. Christie DB, 3rd, Nemec HM, Scott AM, et al. Early outcomes with utilization of tissue plasminogen activator in COVID-19 associated respiratory distress: a series of five cases. J Trauma Acute Care Surg 2020; 89(3): 448–52.

26. Papamichalis P, Papadogoulas A, Katsiafylloudis P, et al. Combination of thrombolytic and immunosuppressive therapy for coronavirus disease 2019: A case report. Int J Infect Dis 2020; 97: 90–3.

27. Poor HD, Ventetuolo CE, Tolbert T, et al. COVID-19 critical illness pathophysiology driven by diffuse pulmonary thrombi and pulmonary endothelial dysfunction responsive to thrombolysis. Clin Transl Med 2020; 10(2): e44.

28. Arachchillage DJ, Stacey A, Akor F, Scotz M, Laffan M. Thrombolysis restores perfusion in COVID 19 hypoxia. Br J Haematol 2020: 10.1111/bjh.17050.

29. Asakura H, Ogawa H. Potential of heparin and nafamostat combination therapy for COVID-19. J Thromb Haemost 2020; 18(6): 1521–2.

30. Doi K, Ikeda M, Hayase N, Moriya K, Morimura N. Nafamostat mesylate treatment in combination with favipiravir for patients critically ill with COVID-19: a case series. Crit Care 2020; 24(1): 392.

31. Thierry AR. Anti-protease treatments targeting plasmin(ogen) and neutrophil elastase may be beneficial in fighting COVID-19. Physiol Rev 2020; 100(4): 1597–8.

32. Oxley TJ, Mocco J, Majidi S, et al. Large-vessel stroke as a presenting feature of COVID-19 in the young. N Engl J Med 2020; 382(20): e60.

33. Chen G, Wu D, Guo W, et al. Clinical and immunological features of severe and moderate coronavirus disease 2019. J Clin Invest 2020; 130(5): 2620–9.

34. Chen T, Wu D, Chen H, et al. Clinical characteristics of 113 deceased patients with coronavirus disease 2019: retrospective study. Bmj 2020; 368: m1091.

35. Feng Y, Ling Y, Bai T, et al. COVID-19 with different severities: a multicenter study of clinical features. Am J Respir Crit Care Med 2020; 201(11): 1380–8.

36. Huang C, Wang Y, Li X, et al. Clinical features of patients infected with 2019 novel coronavirus in Wuhan, China. Lancet 2020; 395(10223): 497–506.

37. Mo P, Xing Y, Xiao Y, et al. Clinical characteristics of refractory COVID-19 pneumonia in Wuhan, China. Clin Infect Dis 2020: 10.1093/cid/ciaa270.

38. Paranjpe I, Russak A, De Freitas JK, et al. Clinical characteristics of hospitalized COVID-19 patients in New York City. medRxiv 2020: 2020.04.19.20062117.

39. Ranucci M, Ballotta A, Di Dedda U, et al. The procoagulant pattern of patients with COVID-19 acute respiratory distress syndrome. J Thromb Haemost 2020; 18(7): 1747–51.

40. Rentsch CT, Kidwai-Khan F, Tate JP, et al. COVID-19 testing, hospital admission, and intensive care among 2,026,227 United States veterans aged 54-75 years. medRxiv 2020: 2020.04.09.20059964.

41. Wang L, He W, Yu X, et al. Coronavirus disease 2019 in elderly patients: characteristics and prognostic factors based on 4-week follow-up. J Infect 2020; 80(6): 639–45.

42. Wang Y, Lu X, Li Y, et al. Clinical course and outcomes of 344 intensive care patients with COVID-19. Am J Respir Crit Care Med 2020; 201(11): 1430–4.

43. Wang Z, Yang B, Li Q, Wen L, Zhang R. Clinical features of 69 cases with coronavirus disease 2019 in Wuhan, China. Clin Infect Dis 2020; 71(15): 769–77.

44. Wu J, Liu J, Zhao X, et al. Clinical characteristics of imported cases of COVID-19 in Jiangsu province: a multicenter descriptive study. Clin Infect Dis 2020; 71(15): 706–12.

45. Yang F, Shi S, Zhu J, Shi J, Dai K, Chen X. Clinical characteristics and outcomes of cancer patients with COVID-19. J Med Virol 2020: 10.1002/jmv.25972.

46. Zhang JJ, Dong X, Cao YY, et al. Clinical characteristics of 140 patients infected with SARS-CoV-2 in Wuhan, China. Allergy 2020; 75(7): 1730–41.

47. Zhang L, Yan X, Fan Q, et al. D-dimer levels on admission to predict in-hospital mortality in patients with Covid-19. J Thromb Haemost 2020; 18(6): 1324–9.

48. Zou Y, Guo H, Zhang Y, et al. Analysis of coagulation parameters in patients with COVID-19 in Shanghai, China. Biosci Trends 2020: 10.5582/bst.2020.03086.

49. Panigada M, Bottino N, Tagliabue P, et al. Hypercoagulability of COVID-19 patients in intensive care unit. A report of thromboelastography findings and other parameters of hemostasis. J Thromb Haemost 2020; 18(7): 1738–42.

50. Su Z, Zhu L, Wu J, Zhao R, Ji HL. Systematic review and meta-analysis of nasal potential difference in hypoxia-induced lung injury. Sci Rep 2016; 6: 30780.

51. Zhao R, Su Z, Wu J, Ji HL. Serious adverse events of cell therapy for respiratory diseases: a systematic review and meta-analysis. Oncotarget 2017; 8(18): 30511–23.

52. Coccheri S. COVID-19: The crucial role of blood coagulation and fibrinolysis. Intern Emerg Med 2020: 1–5.

53. Martín-Rojas RM, Pérez-Rus G, Delgado-Pinos VE, et al. COVID-19 coagulopathy: an in-depth analysis of the coagulation system. Eur J Haematol 2020: 10.1111/ejh.13501.

54. Weisel JW, Litvinov RI. Fibrin formation, structure and properties. Subcell Biochem 2017; 82: 405–56.

55. Takeda Y. Studies of the metabolism and distribution of fibrinogen in healthy men with autologous 125-I-labeled fibrinogen. J Clin Invest 1966; 45(1): 103–11.

56. Favresse J, Lippi G, Roy PM, et al. D-dimer: preanalytical, analytical, postanalytical variables, and clinical applications. Crit Rev Clin Lab Sci 2018; 55(8): 548–77.

57. Rühl H, Berens C, Winterhagen A, Müller J, Oldenburg J, Pötzsch B. Label-free kinetic studies of hemostasis-related biomarkers including D-dimer using autologous serum transfusion. PLoS One 2015; 10(12): e0145012.

58. Olson JD. D-dimer: an overview of hemostasis and fibrinolysis, assays, and clinical applications. Adv Clin Chem 2015; 69: 1–46.

59. Haase C, Joergensen M, Ellervik C, Joergensen MK, Bathum L. Age- and sex-dependent reference intervals for D-dimer: evidence for a marked increase by age. Thromb Res 2013; 132(6): 676–80.

60. Ji HL, Zhao R, Matalon S, Matthay MA. Elevated plasmin(ogen) as a common risk factor for COVID-19 susceptibility. Physiol Rev 2020; 100(3): 1065–75.

61. Johnson ED, Schell JC, Rodgers GM. The D-dimer assay. Am J Hematol 2019; 94(7): 833–9.

62. Nougier C, Benoit R, Simon M, et al. Hypofibrinolytic state and high thrombin generation may play a major role in sars-cov2 associated thrombosis. J Thromb Haemost 2020; 18: 2215–9.

63. Zuo Y, Warnock M, Harbaugh A, et al. Plasma tissue plasminogen activator and plasminogen activator inhibitor-1 in hospitalized COVID-19 patients. medRxiv 2020: 2020.08.29.20184358.

64. Kwaan HC. Coronavirus disease 2019: the role of the fibrinolytic system from transmission to organ injury and sequelae. Semin Thromb Hemost 2020: 10.1055/s-0040-1709996.

65. Bester J, Matshailwe C, Pretorius E. Simultaneous presence of hypercoagulation and increased clot lysis time due to IL-1 β, IL-6 and IL-8. Cytokine 2018; 110: 237–42.

66. Lapic I, Rogic D, Plebani M. Erythrocyte sedimentation rate is associated with severe coronavirus disease 2019 (COVID-19): a pooled analysis. Clin Chem Lab Med 2020; 58(7): 1146–8.

67. Lippi G, Plebani M. Procalcitonin in patients with severe coronavirus disease 2019 (COVID-19): A meta-analysis. Clin Chim Acta 2020; 505: 190–1.

68. Violi F, Ceccarelli G, Cangemi R, et al. Hypoalbuminemia, coagulopathy, and vascular disease in COVID-19. Circ Res 2020; 127(3): 400–1.

69. Semeraro F, Colucci M, Caironi P, et al. Platelet drop and fibrinolytic shutdown in patients with sepsis. Crit Care Med 2018; 46(3): e221–e8.

70. Weiss E, Roux O, Moyer JD, et al. Fibrinolysis resistance: a potential mechanism underlying COVID-19 coagulopathy. Thromb Haemost 2020; 120(9): 1343–5.

71. Henry BM, Benoit SW, Hoehn J, Lippi G, Favaloro EJ, Benoit JL. Circulating plasminogen concentration at admission in patients with coronavirus disease 2019 (COVID-19). Semin Thromb Hemost 2020: 10.1055/s-0040-1715454.

72. Pavoni V, Gianesello L, Pazzi M, Stera C, Meconi T, Frigieri FC. Evaluation of coagulation function by rotation thromboelastometry in critically ill patients with severe COVID-19 pneumonia. J Thromb Thrombolysis 2020; 50(2): 281–6.

73. Tang N, Bai H, Xiong D, Sun Z. Specific coagulation markers may provide more therapeutic targets in COVID-19 patients receiving prophylactic anticoagulant. J Thromb Haemost 2020: 10.1111/jth.14988.

74. Idell S, James KK, Levin EG, et al. Local abnormalities in coagulation and fibrinolytic pathways predispose to alveolar fibrin deposition in the adult respiratory distress syndrome. J Clin Invest 1989; 84(2): 695–705.

75. Idell S. Coagulation, fibrinolysis, and fibrin deposition in acute lung injury. Crit Care Med 2003; 31(4 Suppl): S213–20.

76. Matthay MA, Idell S. Update on acute lung injury and critical care medicine 2009. Am J Respir Crit Care Med 2010; 181(10): 1027–32.

77. Thachil J. All those D-dimers in COVID-19. J Thromb Haemost 2020; 18(8): 2075–6.

78. Hunt BJ, Levi M. Re The source of elevated plasma D-dimer levels in COVID-19 infection. Br J Haematol 2020; 190(3): e133–e4.

79. Wu Y, Wang T, Guo C, et al. Plasminogen improves lung lesions and hypoxemia in patients with COVID-19. Qjm 2020; 113(8): 539–45.

80. Zhao R, Ali G, Nie HG, et al. Plasmin improves blood-gas barrier function in oedematous lungs by cleaving epithelial sodium channels. Br J Pharmacol 2020; 177(13): 3091–106.

81. Liu C, Ma Y, Su Z, et al. Meta-analysis of preclinical studies of fibrinolytic therapy for acute lung injury. Front Immunol 2018; 9: 1898.

82. Schefold JC, Gerber JL, Angehrn MC, et al. Renal function-adjusted D-dimer levels in critically ill patients with suspected thromboembolism. Crit Care Med 2020; 48(4): e270–e6.

83. Robert-Ebadi H, Bertoletti L, Combescure C, Le Gal G, Bounameaux H, Righini M. Effects of impaired renal function on levels and performance of D-dimer in patients with suspected pulmonary embolism. Thromb Haemost 2014; 112(3): 614–20.

84. Cheng A, Hu L, Wang Y, et al. Diagnostic performance of initial blood urea nitrogen combined with D-dimer levels for predicting in-hospital mortality in COVID-19 patients. Int J Antimicrob Agents 2020; 56(3): 106110.

85. Su TH, Kao JH. The clinical manifestations and management of COVID-19-related liver injury. J Formos Med Assoc 2020; 119(6): 1016–8.

86. Nunes Duarte-Neto A, de Almeida Monteiro RA, da Silva LFF, et al. Pulmonary and systemic involvement of COVID-19 assessed by ultrasound-guided minimally invasive autopsy. Histopathology 2020: 10.1111/his.14160.

87. Liu Q, Li C. Predictive value of myoglobin and D-dimer on severe heat stroke: a clinical analysis of 38 patients with severe heat stroke. Chin Crit Care Med 2019; 31(5): 594–7.

88. Simes J, Robledo KP, White HD, et al. D-dimer predicts long-term cause-specific mortality, cardiovascular events, and cancer in patients with stable coronary heart disease: LIPID study. Circulation 2018; 138(7): 712–23.

89. Wu J, Huang J, Zhu G, et al. Elevation of blood glucose level predicts worse outcomes in hospitalized patients with COVID-19: a retrospective cohort study. BMJ Open Diabetes Res Care 2020; 8(1): e001476.

90. Lee TF, Drake SM, Roberts GW, et al. Relative hyperglycemia is an independent determinant of in-hospital mortality in patients with critical illness. Crit Care Med 2020; 48(2): e115–e22.

91. Alzahrani SH, Ajjan RA. Coagulation and fibrinolysis in diabetes. Diab Vasc Dis Res 2010; 7(4): 260–73.

92. Li Q, Cao Y, Chen L, et al. Hematological features of persons with COVID-19. Leukemia 2020; 34(8): 2163–72.

93. Shah S, Shah K, Patel SB, et al. Elevated D-dimer levels are associated with increased risk of mortality in COVID-19: a systematic review and meta-analysis. Cardiol Rev 2020: 10.1097/crd.0000000000000330.

94. Vidali S, Morosetti D, Cossu E, et al. D-dimer as an indicator of prognosis in SARS-CoV-2 infection: a systematic review. ERJ Open Res 2020; 6(2): 00260–2020.

95. Li Q, Chen L, Li Q, et al. Cancer increases risk of in-hospital death from COVID-19 in persons <65 years and those not in complete remission. Leukemia 2020; 34(9): 2384–91.

96. Lima WG, Barra A, Brito JCM, Nizer WSC. D-Dimer serum levels as a biomarker associated for the lethality in patients with coronavirus disease 2019: a meta-analysis. Blood Coagul Fibrinolysis 2020; 31(5): 335–8.

97. Ye W, Chen G, Li X, et al. Dynamic changes of D-dimer and neutrophil-lymphocyte count ratio as prognostic biomarkers in COVID-19. Respir Res 2020; 21(1): 169.

98. Fu J, Kong J, Wang W, et al. The clinical implication of dynamic neutrophil to lymphocyte ratio and D-dimer in COVID-19: A retrospective study in Suzhou China. Thromb Res 2020; 192: 3–8.

99. Li Y, Zhao K, Wei H, et al. Dynamic relationship between D-dimer and COVID-19 severity. Br J Haematol 2020; 190(1): e24–e7.

100. Wang M, Zhang J, Ye D, et al. Time-dependent changes in the clinical characteristics and prognosis of hospitalized COVID-19 patients in Wuhan, China: A retrospective study. Clin Chim Acta 2020; 510: 220–7.

101. Yu B, Li X, Chen J, et al. Evaluation of variation in D-dimer levels among COVID-19 and bacterial pneumonia: a retrospective analysis. J Thromb Thrombolysis 2020: 10.1007/s11239-020-02171-y.

102. Borba MGS, Val FFA, Sampaio VS, et al. Effect of high vs low doses of chloroquine diphosphate as adjunctive therapy for patients hospitalized with severe acute respiratory syndrome coronavirus 2 (SARS-CoV-2) infection: a randomized clinical trial. JAMA Netw Open 2020; 3(4): e208857.

103. Chen N, Zhou M, Dong X, et al. Epidemiological and clinical characteristics of 99 cases of 2019 novel coronavirus pneumonia in Wuhan, China: a descriptive study. Lancet 2020; 395(10223): 507–13.

104. Cui S, Chen S, Li X, Liu S, Wang F. Prevalence of venous thromboembolism in patients with severe novel coronavirus pneumonia. J Thromb Haemost 2020; 18(6): 1421–4.

105. Du RH, Liang LR, Yang CQ, et al. Predictors of mortality for patients with COVID-19 pneumonia caused by SARS-CoV-2: a prospective cohort study. Eur Respir J 2020; 55(5): 2000524.

106. Du Y, Tu L, Zhu P, et al. Clinical features of 85 fatal cases of COVID-19 from Wuhan. A retrospective observational study. Am J Respir Crit Care Med 2020; 201(11): 1372–9.

107. Fogarty H, Townsend L, Ni Cheallaigh C, et al. COVID19 coagulopathy in Caucasian patients. Br J Haematol 2020; 189(6): 1044–9.

108. Han H, Yang L, Liu R, et al. Prominent changes in blood coagulation of patients with SARS-CoV-2 infection. Clin Chem Lab Med 2020; 58(7): 1116–20.

109. Qiu H, Wu J, Hong L, Luo Y, Song Q, Chen D. Clinical and epidemiological features of 36 children with coronavirus disease 2019 (COVID-19) in Zhejiang, China: an observational cohort study. Lancet Infect Dis 2020; 20(6): 689–96.

110. Spiezia L, Boscolo A, Poletto F, et al. COVID-19-related severe hypercoagulability in patients admitted to intensive care unit for acute respiratory failure. Thromb Haemost 2020; 120(6): 998–1000.

111. Tang N, Bai H, Chen X, Gong J, Li D, Sun Z. Anticoagulant treatment is associated with decreased mortality in severe coronavirus disease 2019 patients with coagulopathy. J Thromb Haemost 2020; 18(5): 1094–9.

112. Wu F, Zhao S, Yu B, et al. A new coronavirus associated with human respiratory disease in China. Nature 2020; 579(7798): 265–9.

113. Yang W, Cao Q, Qin L, et al. Clinical characteristics and imaging manifestations ofthe 2019 novel coronavirus disease (COVID-19):A multi-center study in Wenzhou city, Zhejiang, China. J Infect 2020; 80(4): 388–93.

114. Yang X, Yu Y, Xu J, et al. Clinical course and outcomes of critically ill patients with SARS-CoV-2 pneumonia in Wuhan, China: a single-centered, retrospective, observational study. Lancet Respir Med 2020; 8(5): 475–81.

115. Yao Y, Cao J, Wang Q, et al. D-dimer as a biomarker for disease severity and mortality in COVID-19 patients: a case control study. J Intensive Care 2020; 8: 49.

